# Finite element simulation of the pharmacodynamic model for aflibercept and ranibizumab for the treatment of age related macular degeneration

**DOI:** 10.64898/2026.02.05.26345707

**Authors:** Alexander Drobny, Florian T.A. Kretz, Elfriede Friedmann

## Abstract

Age related macular degeneration is known to be one of the major causes of irreversible blindness among the older generation. We present a mathematical model of partial differential equations for the therapy of this disease, which is based on the intravitreal injection of a drug into the vitreous body. For the treatment to work, the drug has to travel past the inner-limiting membrane into the retina and reduce the free vascular endothelial growth factor (VEGF) concentration by binding to at least one of the two binding sites of the VEGF molecule. Therefore, our model consists of two compartments, the vitreous and the retina. In the vitreous we employ four coupled convection-diffusion-reaction equations with an additional coupling to the underlying aqueous humor flow and four coupled diffusion-reaction equations in the retina. The resulting PDE system is solved numerically in a realistic 3D eye geometry. Temporal discretization is based on one-step theta schemes and spatial discretization is done using the Finite Element method. The numerical results are used to demonstrate the therapy concept and to analyze the drug efficacy of aflibercept and ranibizumab. The results show, among other things, that only about 20 % of the drug reaches the retina through the inner-limiting membrane and that 50 % of the VEGF concentration has been rebuilt in the retina after 38.19 days for a single ranibizumab injection.

## Introduction

Age related macular degeneration (AMD) is one of the most common reasons for blindness in the older generation [1]. The typical treatment consists of an intravitreal injection into the vitreous with the aim of decreasing the vascular endothelial growth factor (VEGF) concentration in the retina. After the injection the drug diffuses over time and is being transported by the fluid’s convection in the vitreous. The drug distribution is affected by a number of phenomena: The physiology of the vitreous influences the flow in the vitreous as well as the diffusivity of the drug. The underlying physiology of the healthy vitreous is modeled by a viscoelastic Burgers-type model to include its collagen fibrils and the aqueous humor flow [2, 3]. The aging vitreous naturally liquefies over time or can be replaced during a vitrectomy as a treatment for retinal detachment. This pathology is modeled by the Navier-Stokes equations. In order to reduce the VEGF concentration the drug has to bind to at least one of the two binding sites of a VEGF molecule building a drug-VEGF complex or a drug-VEGF-drug complex [4]. This is included in our model by four coupled convection-diffusion-reaction equations. The binding mechanism used for the reaction terms is based on [5] where a system of ordinary differential equations was analyzed. Our model consists of two compartments, coupled via interface conditions, namely the vitreous and the retina. This gives a better understanding of the processes in the retina which is the main area of interest and allows the modeling of the VEGF production in the retina. For the drug to reach the retina it has to travel past the permeable inner-limiting membrane (ILM) into the retina before clearing through the retinal pigmented epithelium (RPE). In our approach we do not include the anterior chamber as an additional domain since the amount of concentration in the anterior chamber is not of medical interest and since the mass travelling back from the anterior chamber to the vitreous is negligible [5]. The mechanism of anterior clearance is still accounted for through boundary conditions. The elastic structures, specifically the sclera and cornea, are incorporated in [13, 14] leading to a Fluid-Structure Interaction (FSI) problem. It was shown that, given the slow and nearly stationary flow of aqueous humor and the small volume of the injected drug, the elastic sturctures can be disregarded in this analysis. Since the binding constants vary immensely for different drugs we perform numerical simulations for two common drugs, namely aflibercept and ranibizumab and analyze their efficacy by studying the behavior of the average concentration in the vitreous as well as in the retina over time.

PDE models for drug therapy have previously been studied in the literature. Most studies consider only the drug distribution in the vitreous, modeled by one convection-diffusion equation, and use Darcy’s law for the flow equations (e.g. [6, 7]). In [8] this model was used but further therapeutic approaches have been analyzed using finite element simulations: the influence of gravity and diffusion coefficient on drug distribution and the optimal injection position and angle were discussed. The Virtual Eye [11, 12], which we also use here, was presented there, in particular the geometry of the vitreous, and different functionals were developed to monitor the amount of drug remaining in the vitreous or operating at the macula at a given time. The influence of collagen fibers in the vitreous on drug distribution was considered by anisotropic diffusion. Here, we go into pharmacological details and consider the binding and complex formation, and the viscoelastic structure of the vitreous is also considered by using Burger’s instead of Darcy’s model. And we include the retinal compartment where the disease starts developing.

The advantage of our approach is that we additionally gain information about the VEGF concentration, the concentrations of the two complexes and the amount of concentration in the retina. Furthermore the fluid modeling via the viscoelastic Burgers-type model and the Navier-Stokes equations allows an analysis of the influence of the viscoelasticity of a healthy vitreous and of the water-like vitreous obtained from a vitrectomy on the drug therapy. Finally our model also includes the VEGF production in the retina and can be varied depending on the severity of the disease. In [9] the authors studied a simplified two concentration model, while the four concentration model in one compartment was studied in [10]. The geometry for the vitreous is based on [11, 12].

In Section 1.1 we state the equations for the fluid flow in the vitreous, namely the Burgers-type model for the healthy vitreous and the Navier-Stokes equations for the aging or phathological (liquified) vitreous. Furthermore we present the equations for the chemical processes in the vitreous and in the retina as well as the interface, boundary and initial conditions. In Section 1.2 we derive the variational formulations of the previously stated models and summarize the temporal discretization, the spatial discretization via finite elements and the Gateaux derivatives necessary for Newton’s method. In Section 2 we present the numerical results. In Section 3 we state our conclusion.

## 1. Mathematical modeling and numerical discretization

### 1.1 Mathematical Modeling

In this section we introduce the two-compartment model for the treatment of AMD. The fluid flow in the vitreous obtained from the Burgers-type or the Navier-Stokes equations. This convection enters into the convection-diffusion-reaction equations in the vitreous which are coupled via an interface to a diffusion-reaction system for the chemical processes in the retina. In this work we consider a stationary inflow at the hyaloid membrane into the vitreous. With the model and the numerical implementation it is also possible to study the influence of time-dependent saccadic eye movements on the velocity and the effect of the viscoelasticity of the vitreous. In [13, 14] a time-depended fluid-structure problem with this fluid model was analyzed.

The computational domain Ω ⊂ ℝ^3^ consists of the vitreous Ω_vit_ and the retina Ω_ret_ and is given by Ω = Ω_vit_ ∪ Γ_ILM_ ∪ Ω_ret_. The two domains do not overlap Ω_vit_ ∩Ω_ret_ = ∅and the common interface is denoted by Γ_ILM_ = ∂Ω_vit_ ∩∂Ω_ret_ which corresponds to the inner limiting membrane. Fig 1 shows the domain sliced in the middle for visualization purposes. The boundary Γ consists of the boundary of the lens, the hyaloid membrane, and the retinal pigmented epithelium:

**Fig 1:**
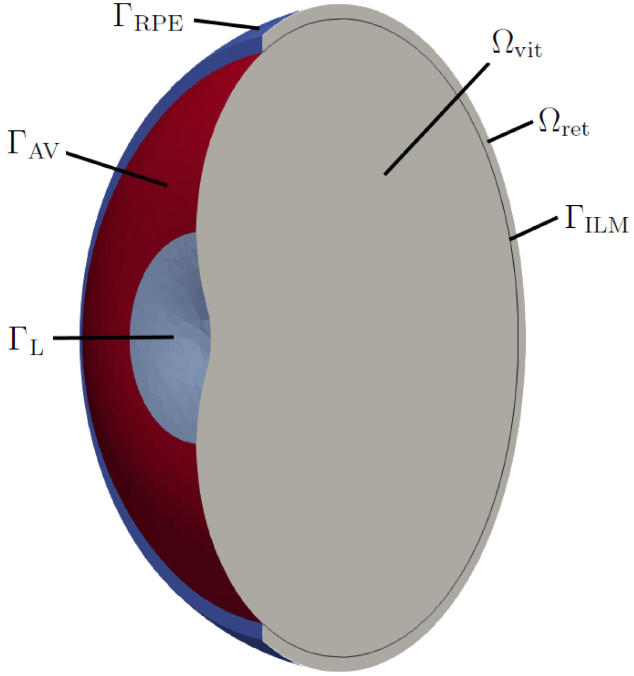
Sketch of the computational domain Ω sliced in the middle for visualization purposes.

Γ = Γ_L_ ∪ Γ_AV_ ∪ Γ_RPE_. Finally let *I* = (0, *T*], *T >* 0 be the time interval and *t* ∈ *I*.

#### 1.1.1 Fluid equations

We start by stating the fluid equations for the vitreous Ω_vit_. For a healthy vitreous we use a viscoelastic Burgers-type model [15]. In recent experiments [2, 3] it has been shown to be capable of capturing the viscoelastic mechanisms of the vitreous. The Burgers-type model is given by

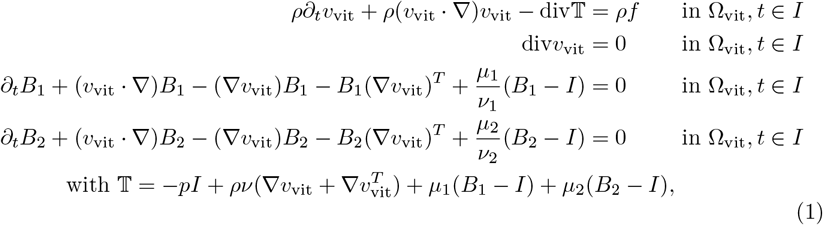

where *ρ* is the density, *ν* the viscosity, *v*_vit_ the velocity, *p* the pressure, *B*_1_ and *B*_2_ tensor-valued unknowns for the viscoelasticity and 𝕋 the Cauchy stress tensor. The liquified vitreous which can be a result of a vitrectomy or a liquification of the vitreous over time is modeled by the Navier-Stokes equations which are obtained from (1) by dropping the terms involving *B*_1_ and *B*_2_. The parameters can be found in Table 1.

**Table 1.** Material parameters for the fluid.

| Parameter | Units | Value | Computation | Source |
| --- | --- | --- | --- | --- |
| $\rho_f$ | kg/m<sup>3</sup> | $1.0053 - 1.0089 \cdot 10^3$ | $1.007 \cdot 10^3$ | [19] |
| $\mu_1$ | Pa | $1.07 \cdot 10^{-1}$ | $1.07 \cdot 10^{-1}$ | [3] |
| $\nu_1$ | Pa · s | 9200.28 | 9200.28 | [3] |
| $\mu_2$ | Pa | $6.99 \cdot 10^{-2}$ | $6.99 \cdot 10^{-2}$ | [3] |
| $\nu_2$ | Pa · s | 4.17 | 4.17 | [3] |
| $\nu$ (healthy) | Pa · s | $8.53 \cdot 10^{-4}$ | $8.53 \cdot 10^{-4}$ | [3] |
| $\nu$ (vitrectomized) | Pa · s | $7.322 \cdot 10^{-7}$ | $7.322 \cdot 10^{-7}$ | [20] |

As boundary conditions we have a parabolic inflow condition at the hyaloid membrane Γ_AV_, the aqueous flow being secreted at the ciliary body, a zero Dirichlet condition on the lens and an outflow condition at the ILM:

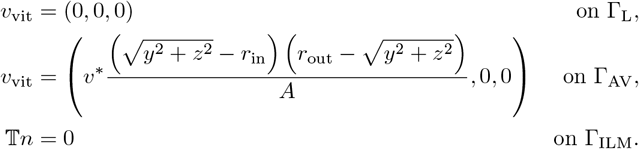

The inflow velocity is approximated using the area of the hyaloid membrane, which is estimated to be 2.4 cm^2^ [16] and the aqueous humour production at the ciliary body which is around around 2.5 *µ*l*/*min [17]. The inner and outer radii of the inflow area are denoted by *r*_in_ and *r*_out_, respectively. The velocity is modeled as a parabolic inflow with a mean inflow velocity at the hyaloid membrane of 8.92 · 10^−9^ m*/*s and a maximum velocity of 1.39 · 10^−8^ m*/*s and *v*^∗^ = 4.42 · 10^−7^m*/*s (see [13] for details).

#### 1.1.2 Chemical processes

The model for the chemical processes consists of a system of four coupled convection-diffusion-reaction equations for the vitreous. The convection is calculated from the Burgers-equations or the Navier-Stokes equations, while the reaction terms model the binding and dissociation of the chemicals [5]. In this work we neglect the flow in the retina due to its small velocity and therefore little impact on the drug and obtain a system of four coupled diffusion-reaction equations with an additional term for the VEGF production in comparison to the vitreous model. Then the model reads:

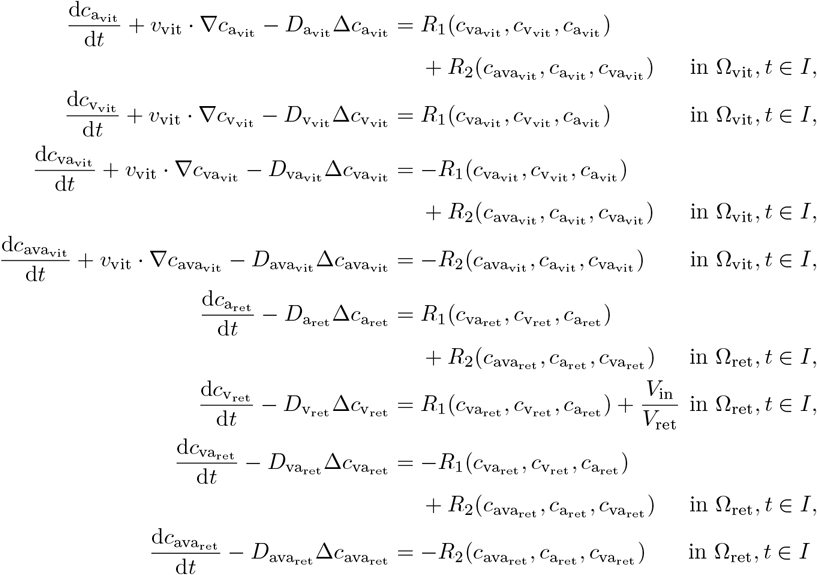

with

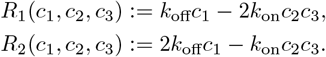

The boundary conditions are of Neumann type and model the permeability of the corresponding membrane. For the definition of the boundary conditions we refer to [13]. The parameters can be found in Table 2.

**Table 2.** Binding parameters for ranibizumab and aflibercept.

| Parameter | Value | Unit | Description | Source |
| --- | --- | --- | --- | --- |
| $k_{\text{on}}$ (aflibercept) | $3.02 \cdot 10^{-7}$ | $\text{pM}^{-1} \text{s}^{-1}$ | binding on rate | [18] |
| $k_{\text{off}}$ (aflibercept) | $280.2 \cdot 10^{-5}$ | $\text{s}^{-1}$ | binding off rate | [18] |
| $k_{\text{on}}$ (ranibizumab) | $5.82 \cdot 10^{-8}$ | $\text{pM}^{-1} \text{s}^{-1}$ | binding on rate | [18] |
| $k_{\text{off}}$ (ranibizumab) | $0.39 \cdot 10^{-5}$ | $\text{s}^{-1}$ | binding off rate | [18] |
| $k_{\text{on}}$ (ranibizumab) | $5.26 \cdot 10^{-10}$ | $\text{pM}^{-1} \text{s}^{-1}$ | binding on rate | [5] |
| $k_{\text{off}}$ (ranibizumab) | $1 \cdot 10^{-5}$ | $\text{s}^{-1}$ | binding off rate | [5] |
| $V_{\text{in}}$ | 18.5 | fmol/day | VEGF production rate | [5] |

Finally we state the initial conditions. The initial conditions model the disease dependent VEGF concentration at the beginning 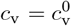 and the injection of the drug. The drug is either assumed to be uniformly distributed in the vitreous or modeled as an injection in a small ball with volume 0.05 ml. To make the initial condition for the second case continuous we use 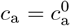 with

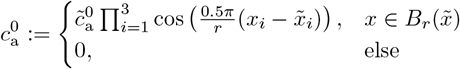

with a ball 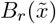 with radius *r* = 2.285 mm and center.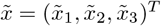 For a uniform injection of ranibizumab and aflibercept, which we refer to in the case when the drug is already uniformly distributed in the vitreous after injection, we have

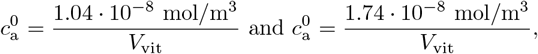

respectively. Here *V*_vit_ = 3.65 10^−6^ m^3^ denotes the volume of the vitreous. For an injection in a small ball we have for example for ranibizumab 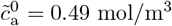. The initial concentration for the two complexes are zero: *c*_va_ = 0, *c*_ava_ = 0.

### 1.2 Numerical Discretization

The variational formulation 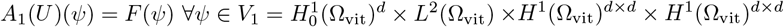 for the Burgers-type model (1) is given by:

**Problem 1** *Find* 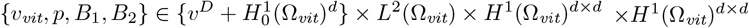 *such that the initial data v*(0) = *v*^0^, *B*_1_(0) = *I, B*_2_(0) = *I are satisfied and for almost all time steps t* ∈ *I it holds:*

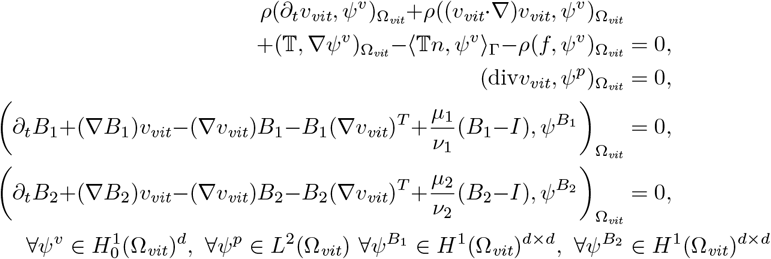

*with the Cauchy stress tensor*

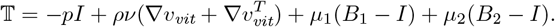

Next we state the variational formulation for the chemical processes,

*A*_2_(*U, ψ*) = *F* (*ψ*) ∀*ψ* ∈*V*_2_ = *H*^1^(Ω_vit_) × *H*^1^(Ω_vit_) × *H*^1^(Ω_vit_) × *H*^1^(Ω_vit_) × *H*^1^(Ω_ret_) × *H*^1^(Ω_ret_) × *H*^1^(Ω_ret_) × *H*^1^(Ω_ret_). Here *v*_vit_ is the velocity obtained from the Burgers or Navier-Stokes equations and is assumed to be sufficiently regular.

**Problem 2** *Find* 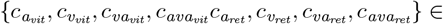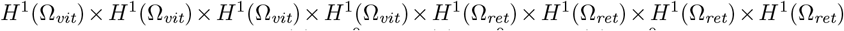 *such that the initial data* 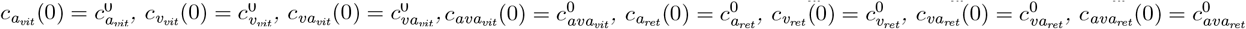 *are satisfied and for almost all time steps t* ∈ *I it holds:*

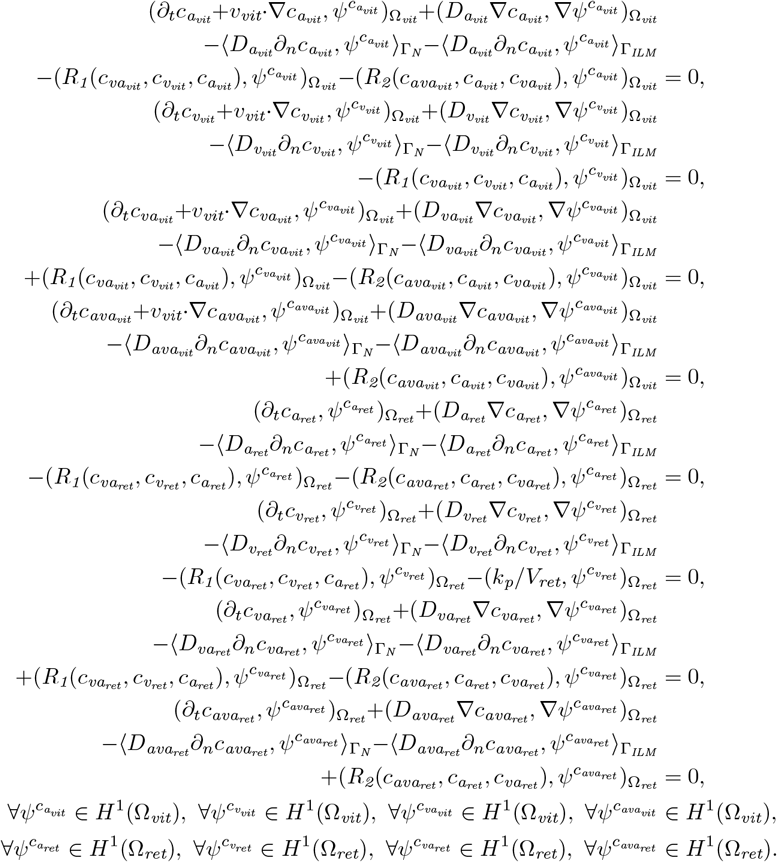

In the following we summarize the numerical discretization of the weak formulations from Problem 1 and Problem 2. For the temporal discretization we split the time interval *I* = [0, *T*] into (here uniform) discrete time steps

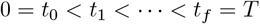

with *k* := *t*_*n*_ − *t*_*n*−1_ (for nonuniform discretization one can use *k*_*n*_ := *t*_*n*_ − *t*_*n*−1_). Temporal discretization is done using one-step theta schemes.

For the spatial discretization we use quadratic Lagrangian finite elements *Q*^2^ for the velocity and the viscoelastic tensors and a linear discontinuous ansatz *P* ^1,dc^ for the pressure. The four concentrations are approximated using linear Lagrangian finite elements *Q*^1^. Since both problems are nonlinear, we use Newton’s method for the linearization. For a detailed derivation and the corresponding Gateaux derivatives we refer to [13].

## 2 Results

The numerical simulations in this work are implemented using the finite element library deal.ii [21]. For convergence studies and a more detailed analysis of the numerical results we refer to [13].

Fig 2 shows the mesh for the two compartment model. It consists of a connected mesh of the vitreous with 43248 cells and the retina with 18768 cells. The colors indicate the various boundaries already described in Section 1.1.

**Fig 2:**
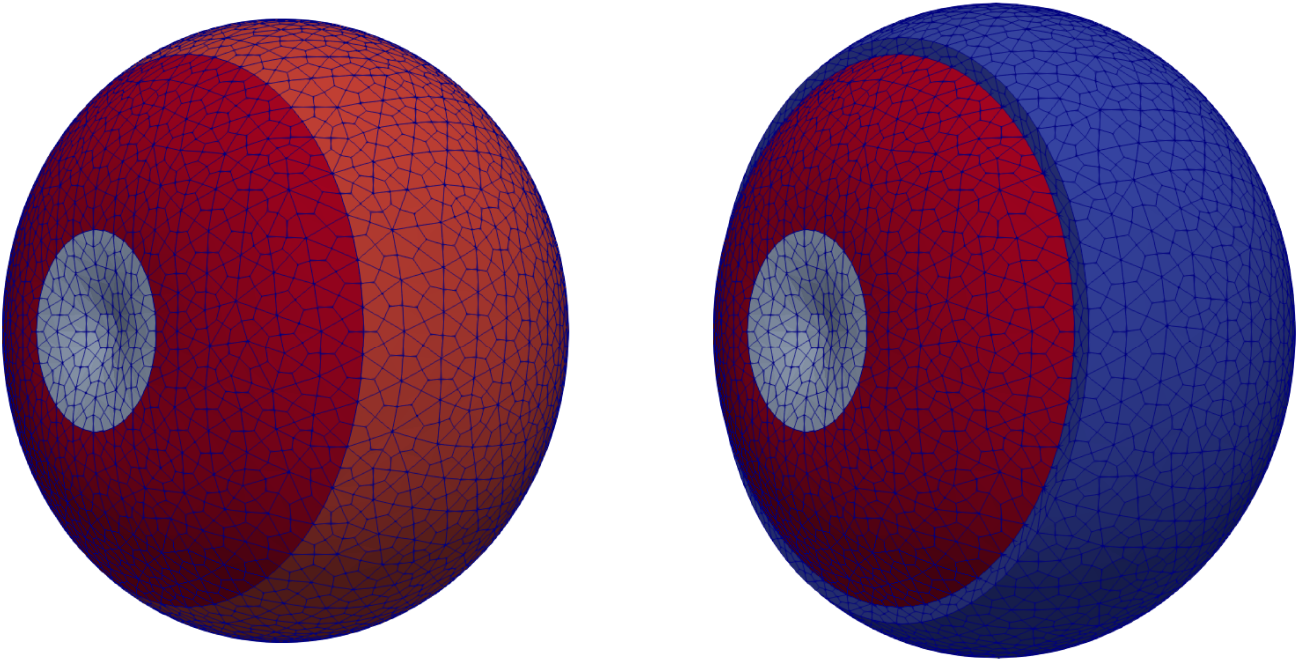
Mesh constructed using Gmsh [22], left: vitreous (gray: boundary to the lens, dark red: inflow boundary (hyaloid membrane), light red: boundary towards retina), right: vitreous and retina (blue).

Fig 3 shows the velocity in the vitreous. The mean velocity is 7.25 · 10^−9^ m*/*s. Due to the rather slow inflow velocity the flow has a stationary character and thus, the influence of the viscoelasticity in the Burgers-type model is small. The model and implementation can be used in the future to study molecular aspects of the drug-vitreous interactions or saccadic eye movements where the velocity is faster and the impact of the viscoelasticity might be much higher due to the instationarity of the movement. Another application involving the effect of the viscoelasticity can be found in [23] by the authors, namely instationary fluid-structure interaction problems in the eye. There the viscoelasticity has a big impact on the stress in the vitreous.

**Fig 3:**
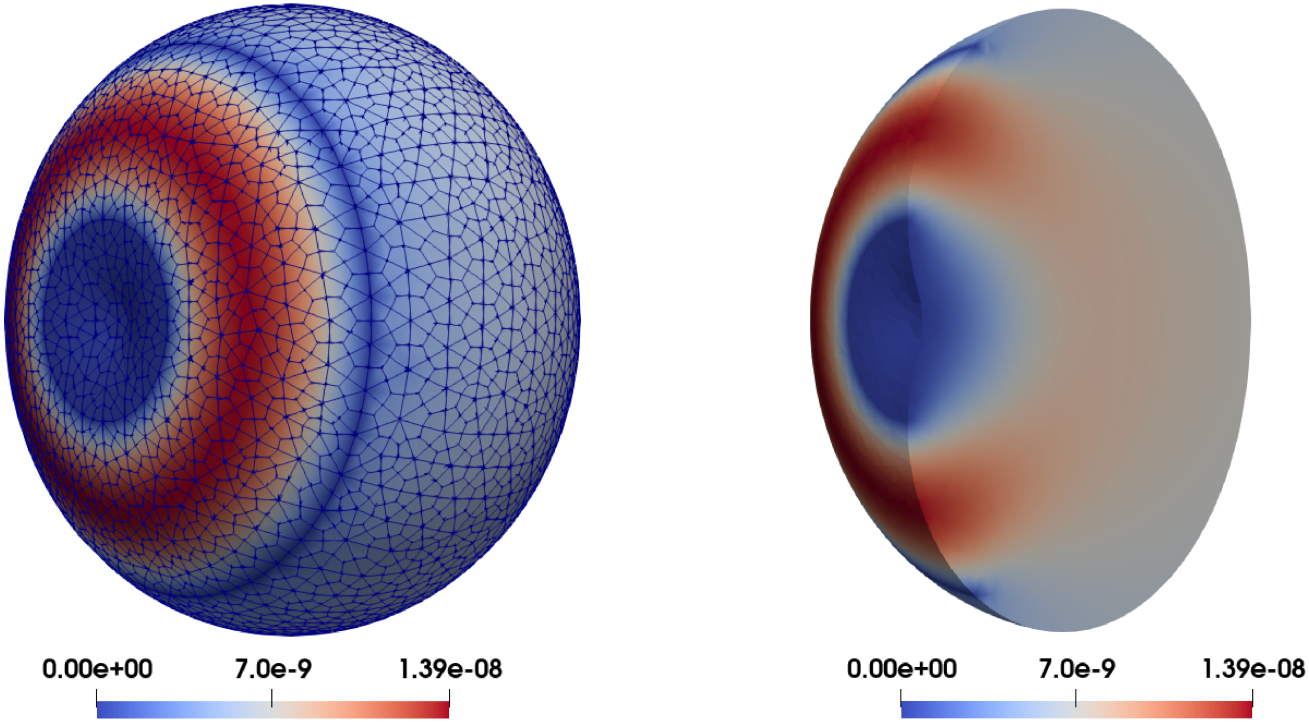
Velocity magnitude in the vitreous in m*/*s for the entire vitreous on the left and for the vitreous cut in half for visualization purposes on the right.

In the following we assume that the VEGF production is space-dependent and centered in the area of the macula. Fig 4 shows the numerical solution for one ranibizumab injection after 0.5, one and 4.5 days. The drug distributes in the vitreous over time and slowly reaches the retina. Since the hyaloid membrane is very permeable the concentration can clear into the anterior part of the eye. Therefore the concentration is lowest in this area. In the region of the ILM the concentration is higher due to the smaller permeability of the membrane and reaches its highest value the furthest away from the hyaloid membrane. Due to the binding mechanism the VEGF concentration is lowest in the area where the drug concentration is high. Furthermore we can see that the VEGF concentration in the vitreous is comparably small at all three times. In the retina the concentration is only reduced after about one day since the drug has to travel through the ILM before reaching the retina. The concentration of the drug-VEGF complex is comparably small due to the fast binding at both VEGF sites resulting in a high drug-VEGF-drug complex concentration. This concentration distributes over time and clears through the RPE and the hyaloid membrane. Fig 5 shows the corresponding results for aflibercept. The distribution behavior of the concentrations is very similar to the previous injection case with ranibizumab, only the level of the concentrations differs. At the posterior retina (around the macula) 5.5*e* − 03 mol/m^3^ of aflibercept can be localized compared to 2.7*e* − 03 mol/m^3^ of ranibizumab. The higher amount of aflibercept near the retina causes a significantly lower VEGF concentration in the retina, 2.7*e* − 11 mol/m^3^ instead of 1.4*e* − 08 mol/m^3^ for the ranibizumab treatment, demonstrating the efficacy of the aflibercept treatment.

**Fig 4:**
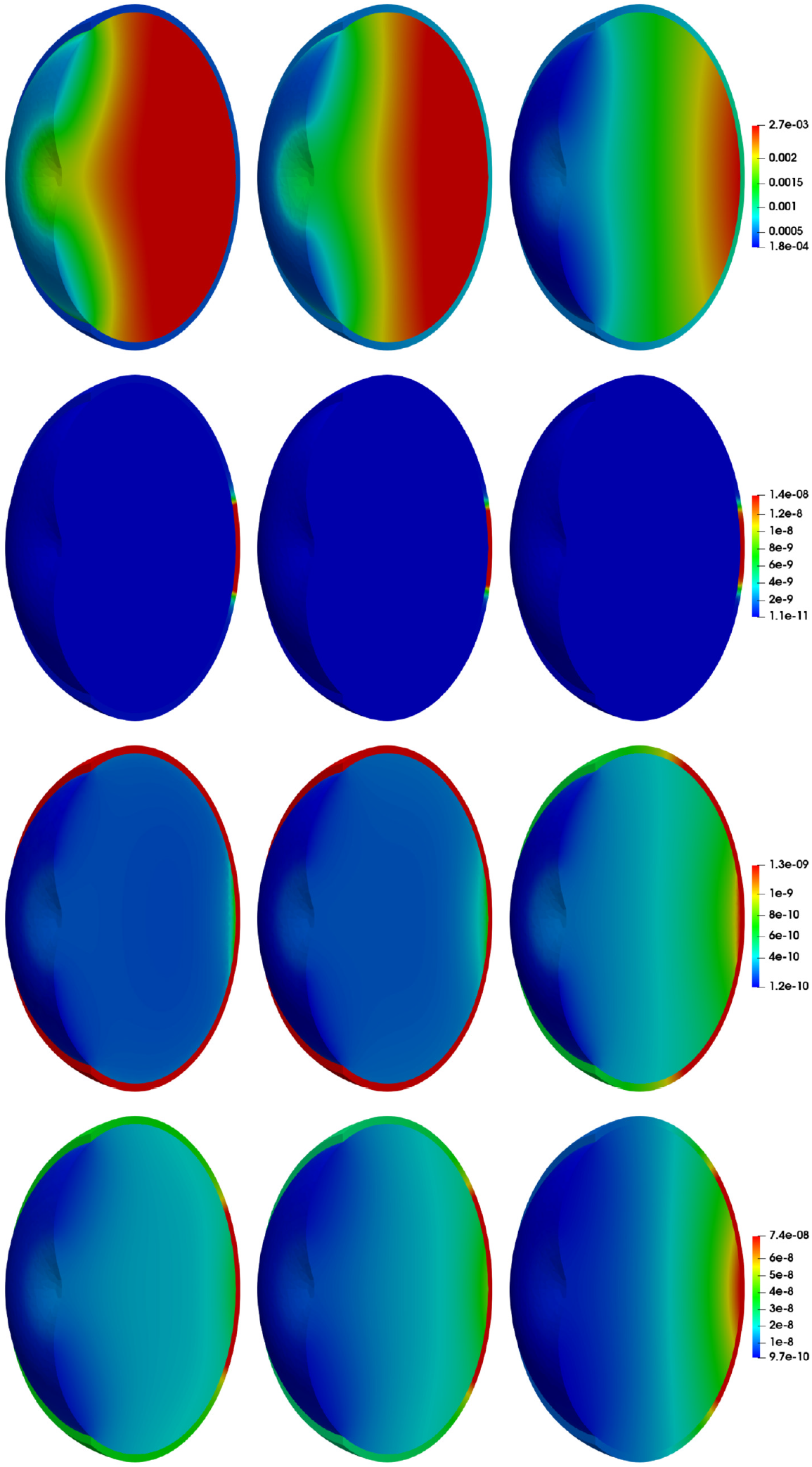
Results for a uniform drug injection for ranibizumab. First column: solution for ranibizumab after 0.5 days, second column: solution after one day, third column: solution after 4.5 days, first row: *c*_a_, second row: *c*_v_, third row: *c*_va_, fourth row: *c*_ava_ . In each row the same scaling is used. The concentrations are in mol*/*m^3^.

**Fig 5:**
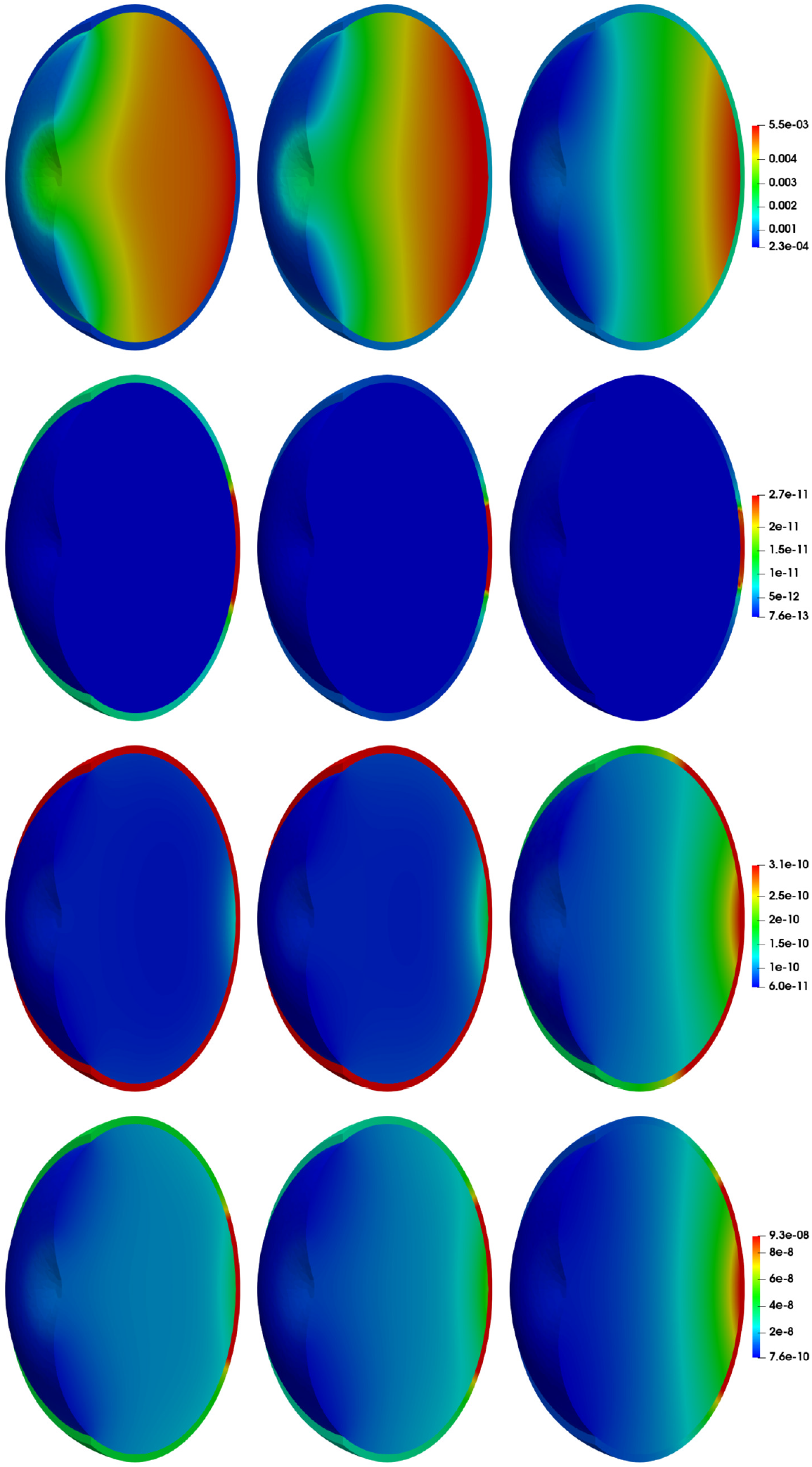
Results for a uniform drug injection for aflibercept. First column: solution after 0.5 days, second column: solution after one day, third column: solution after 4.5 days, first row: *c*_a_, second row: *c*_v_, third row: *c*_va_, fourth row: *c*_ava_ . In each row the same scaling is used. The concentrations are in mol*/*m^3^.

Fig 6 shows the average concentration in the retina and the vitreous for aflibercept and ranibizumab. We additionally consider a different set of binding parameters for ranibizumab (see Table 2). The difference in *k*_on_ is two orders of magnitude and *k*_off_ is similar (factor 2.5). The difference in drug concentration for aflibercept comes from administering a different amount in the intravitreal injection. The half-life time is 5.6 days for aflibercept and 4.5 days for ranibizumab. This difference emerges mainly due to the smaller diffusion coefficient for aflibercept. The VEGF concentration initially decreases faster for the parameters from [18].

**Fig 6:**
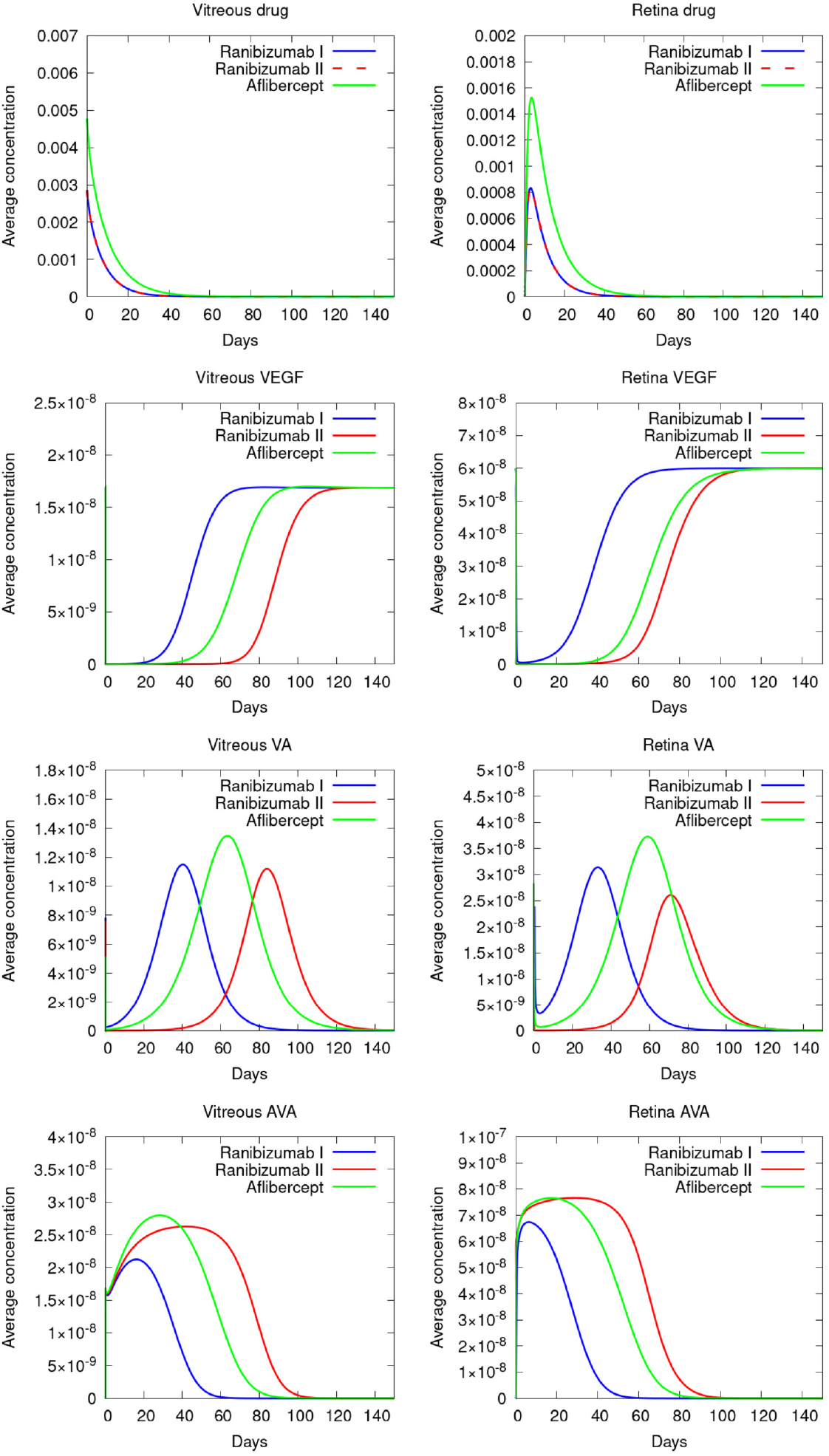
Comparison of the average concentrations over time in mol*/*m^3^ in the vitreous and the retina for two sets of binding parameters for ranibizumab and aflibercept.

As it is expected the drug slowly decreases in the vitreous mainly due to the clearance to the anterior part of the eye and due to the transfer into the retina, where the concentration of the drug increases for the first three days before slowly clearing through the RPE. The last column shows the fast increase in drug-VEGF-drug complexes which slowly clear out of the retina and vitreous in addition to dissociating into drug-VEGF complexes.

Furthermore the VEGF concentration increases much earlier over time for the set of parameters from [5]. This is mostly due to the difference in binding parameters: The drug can bind faster to the continuously produced VEGF in the other two cases. Furthermore the disintegration of the complexes is fastest for aflibercept, while the values are similar for the two ranibizumab cases (see Table 2).

Therefore it seems like the first set of parameters for ranibizumab performs worse than aflibercept since the VEGF concentration starts increasing earlier than in the case of aflibercept. On the other hand the second set of parameters for ranibizumab performs better since the VEGF concentration increases later in the retina than for aflibercept. Differences in the results depending on which set of parameters is used have to be expected since the experimental methods can vary for different studies [24] and the data obtained can be very different depending on the experimental setup [18] yielding big differences in the binding parameters reported in the literature. Nevertheless the simulations can give a better understanding of the drug therapy.

Next we analyze the influence of the location of the drug injection. To this end we additionally consider a drug injection in the center of the vitreous and an injection location at a different position, shifted horizontally towards the hyaloid membrane and vertically downward. We refer to this situation as the shifted injection. The drug distribution of the shifted case can be seen in Fig 7.

**Fig 7:**
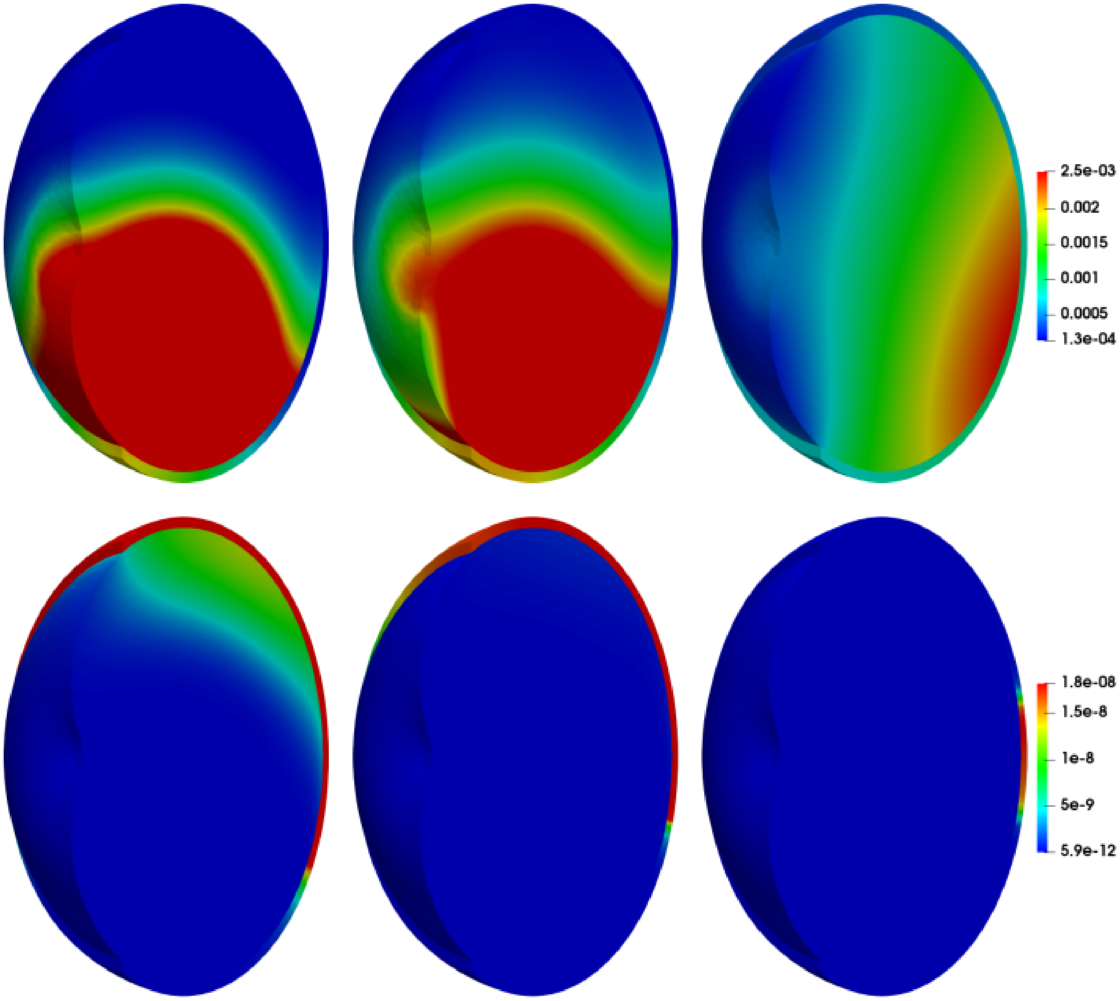
Drug and VEGF concentration after 0.5 days, one day and 4.5 days for the injection in a small ball where the center of the ball is shifted.

Fig 8 shows the corresponding average concentrations over time. We first discuss the results for the first two cases, the uniform injection and the injection in a small ball. The half-life time for the injection in a small ball is slightly longer: For the uniform injection the half-life time is 4.5 days and for the injection in a small ball the half-life time is 5.5 days. The figure also shows that after about 30 days there is only a small amount of the drug left in the vitreous and in the retina. In detail after 30 days for the uniform drug injection there is only 2.30 % and for the injection in a small ball only 2.60 % of the initial drug amount left in the vitreous and in the retina together. Furthermore the highest drug concentration in the retina is reached after 2.9 days for the uniform injection and after 3.7 days for the drug injection in a ball.

**Fig 8:**
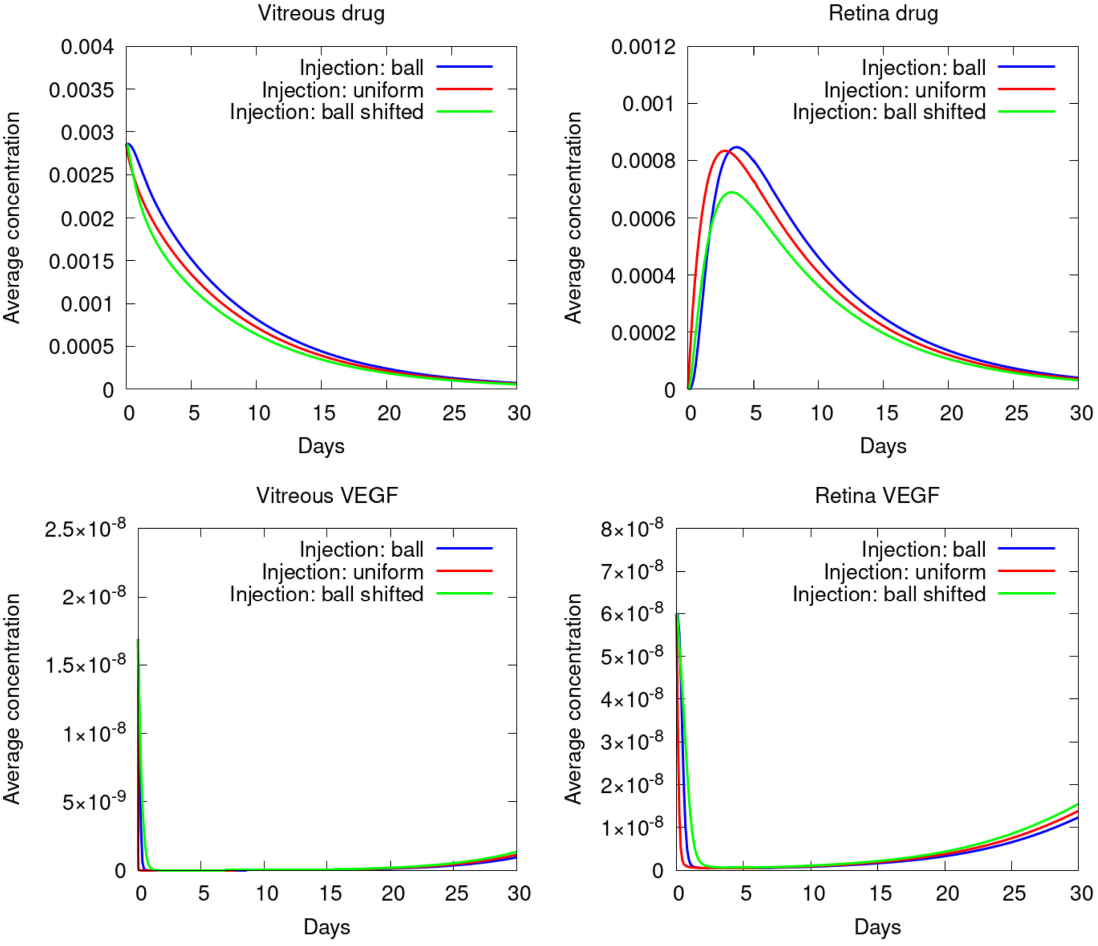
Comparison of the average concentrations over time in mol*/*m^3^ in the vitreous and the retina for an injection in a small ball, an injection where the center of the ball is shifted and a uniform drug distribution at the beginning.

The figure also shows that the amount of VEGF is reduced very fast in both domains. In the case of a uniform drug injection the VEGF is reduced slightly faster since the drug is already present in all areas of the vitreous in the beginning. The VEGF concentration after 30 days is rising again due to the constant VEGF production in the retina, the dissociation of the complexes over time and since there is almost no drug left which can bind to the VEGF. For the uniform injection case the VEGF has reached 50 % of the initial VEGF concentration in the retina after 38.19 days and 90 % after 54.51 days. This shows the necessity of multiple injections stretched over a long period of time. After roughly 75 days the average VEGF concentration has reached its long-term stationary solution in both compartments. Note, that the results refer to constant physiological processes and do not involve any changes in time. In case of interest, changes must be implemented in the model.

Finally, we consider the third case, where a different injection location is chosen, which we called shifted injection. As can be seen in Fig 7, the majority of the drug is concentrated in the lower half of the vitreous and retina due to the shifted injection location. Note, that as shown in [8] the orientation of the eye and gravity has not a big influence, the visible difference in concentration distribution comes from diffusion. The injected drug is no longer in the center of the vitreous, it is located in the lower half of the vitreous and closer to the hyaloid membrane. It diffuses through the vitreous, initially reaching the hyaloid membrane on one side and the lower part of the retina on the oher side. Therefore, there is more VEGF present in the top half of the vitreous and retina than in the lower half. With 3.6 days the half-life time of the drug is considerably shorter than in the other two cases. Since the injection position is closer to the highly permeable hyaloid membrane this has to be expected. As a result less drug reaches the retina (see Figs 8 and 9). The half life time for the injection in a ball and the uniform injection is longer by 25.00 % and 52.78 %, respectively in comparison to the shifted injection. Nevertheless, the results show that this does not severely hamper the VEGF reduction: 50 % of the initial VEGF concentration is reached after 37.38 days. The time when 50 % is reached for the uniform injection is 2.17 % longer with 38.19 days and for the injection in a small ball it is 5.27 % longer with 39.35 days. In the following we restrict the considerations to the first two cases.

**Fig 9:**
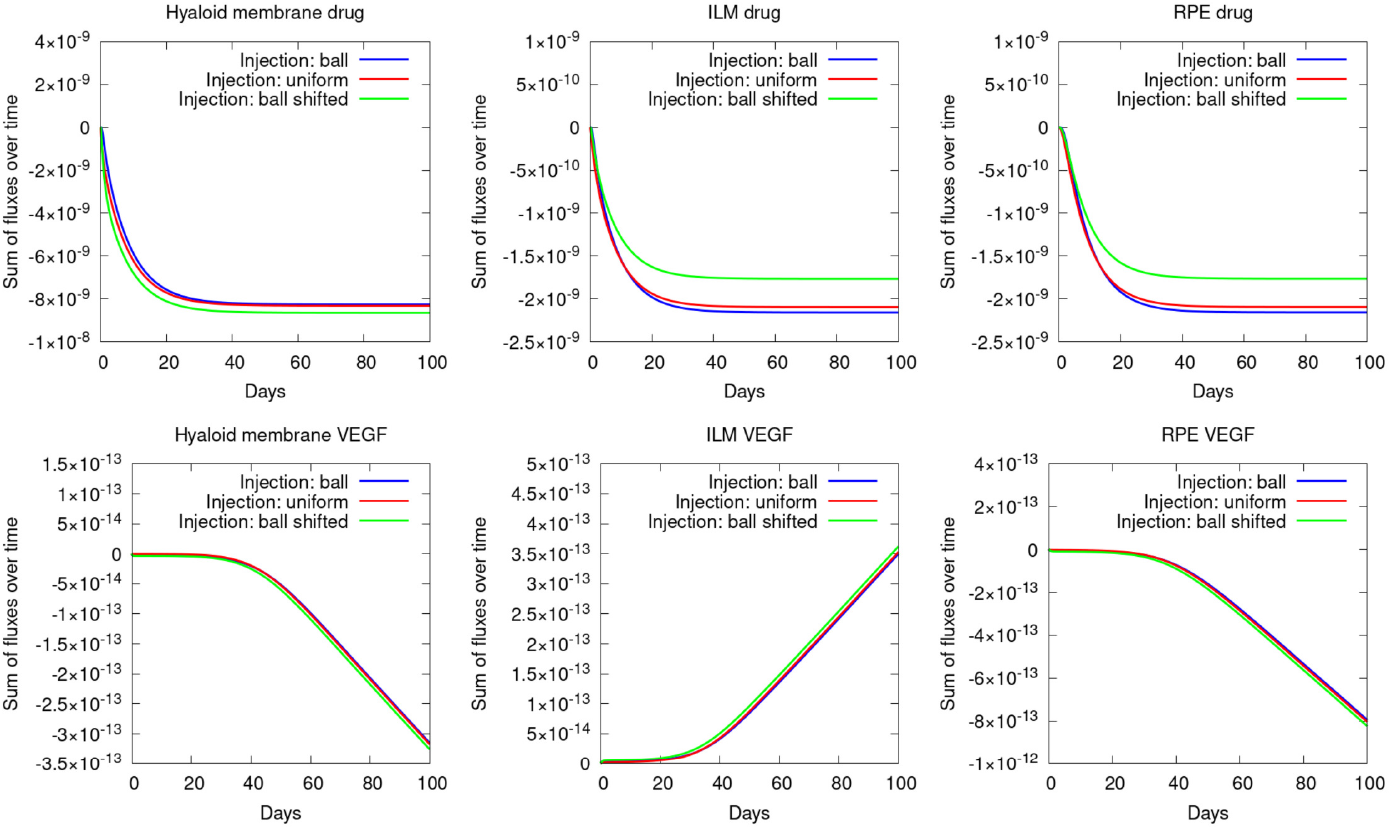
Comparison of the accumulated fluxes over time in mol for an injection in a small ball, an injection where the center of the ball is shifted and a uniform drug distribution at the beginning.

Fig 9 shows the corresponding accumulated fluxes over time at the hyaloid membrane, the ILM and the RPE. The figure shows that a large fraction of the drug clears through the hyaloid membrane into the anterior chamber and therefore only a small fraction reaches the retina through the ILM. Of this fraction almost everything clears through the RPE over time. In detail for a uniform drug injection in the first 50 days 8.3053· 10^−9^ mol has cleared through the hyaloid membrane and 2.0924 ·10^−9^ mol through the ILM. Over time free VEGF which is produced in the retina partially travels into the vitreous.

Next we analyze the influence of convection. Fig 11 shows the average concentrations over time for a uniform drug injection of ranibizumab with convection and without convection. The average drug concentration is higher in both compartments for the case with convection. This can also be seen in Fig 10 which shows the drug concentration at the same times with the same colour scale as in Fig 4. These differences can be expected since the flow direction is from the hyaloid membrane towards the ILM. Therefore the flow decreases the flux through the hyaloid membrane.

**Fig 10:**
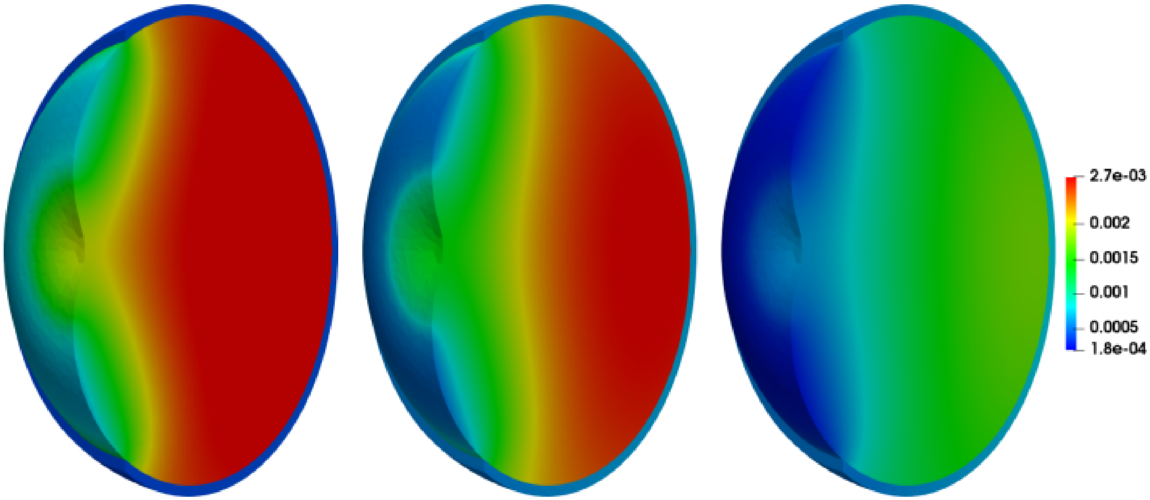
Results for a uniform drug injection for ranibizumab without convection of the fluid. Left: solution for ranibizumab after 0.5 days, middle: solution after one day, right: solution after 4.5 days. The concentrations are in mol*/*m^3^.

**Fig 11:**
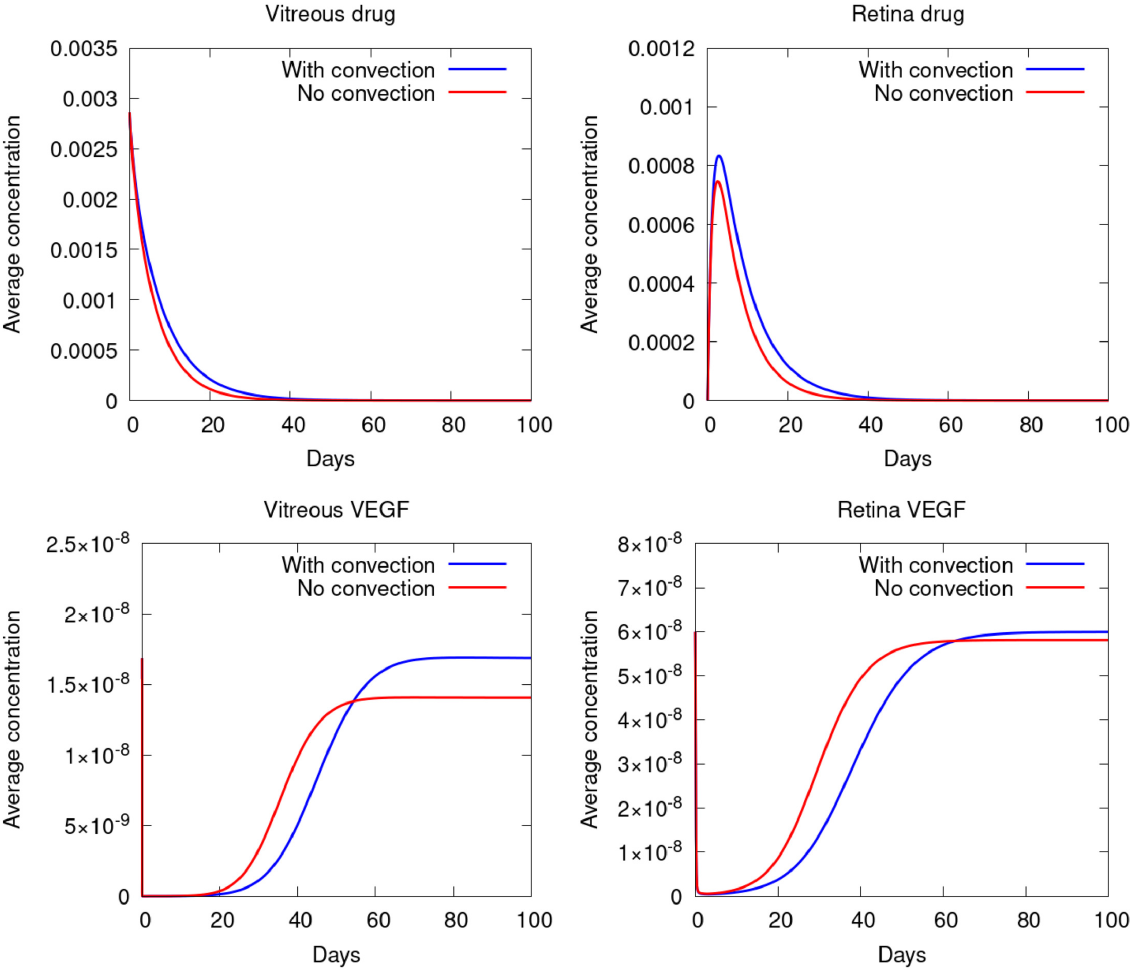
Comparison of the average concentrations over time in mol*/*m^3^ in the vitreous and the retina with and without convection.

Fig 12 shows a comparison of different administration procedures. The first case is for two separate injections 30 days apart, while the second case uses one injection in the beginning with a doubled dose. The third case is the previously studied procedure with one injection and a single dose. The results show that the VEGF concentration in the retina increases much earlier in the case with a doubled dose in comparison to two injections. This result implies that separate regular injections might yield better long-term results for the VEGF reduction. Furthermore too high drug concentrations can be toxic. On the other hand it is known that intravitreal injections might damage the vitreous such that too many injections are not feasible either. Furthermore the figure shows that a double dose of 1 mg does not significantly improve the AMD treatment in comparison to one dose of 0.5 mg: After 50 days the VEGF concentration in the retina for a single dose is 2.19 · 10^−14^ mol and for a double dose 1.83 · 10^−14^ mol. Therefore the improvement of a double dose is rather small. At the same time for two separate injections of 0.5 mg the VEGF concentration is more than ten times smaller with 1.60 · 10^−15^ mol.

**Fig 12:**
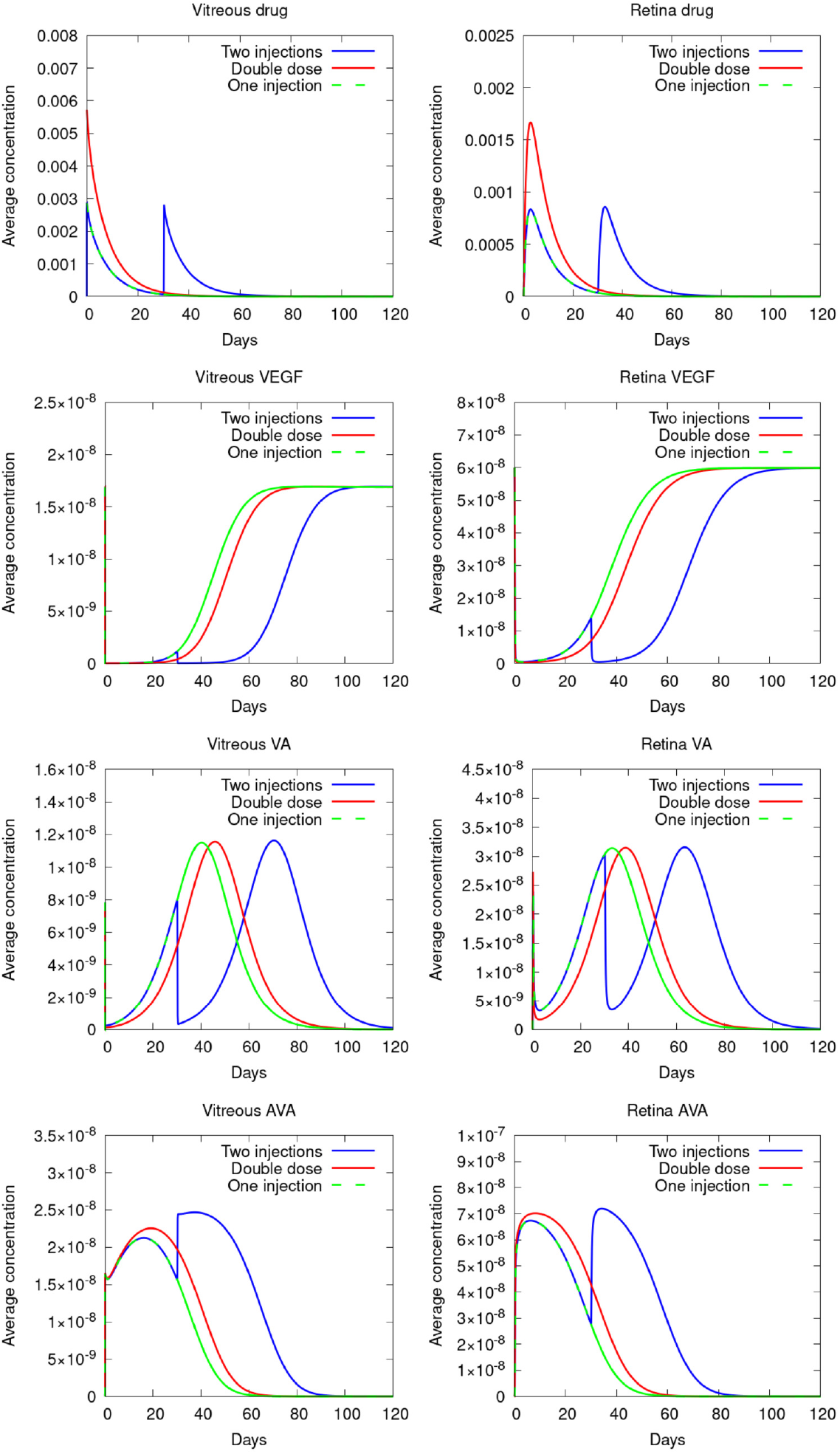
Comparison of the average concentrations over time in mol*/*m^3^ in the vitreous and the retina for two injections spread 30 days apart, an injection with twice the drug amount and a single dose of 0.5 mg.

On the other hand higher doses could be used to increase the time between injections and thus decrease the total amount of necessary injections. Recently, in [25] it has been shown that an 8 mg dose of aflibercept showed efficacy and safety with extended dosing intervals. Fig 13 shows a similar simulation with 2 mg, 4 mg and 8 mg aflibercept. It can be seen that an increased dose extends the effectivity of the drug injection. For example 50 % of the initial VEGF amount which was present in the retina before the injection, is reached after approximately 81, 88 and 95 days for 2 mg, 4 mg and 8 mg, respectively, indicating that higher doses could be used to decrease the amount of necessary injections and increase the time between injections.

**Fig 13:**
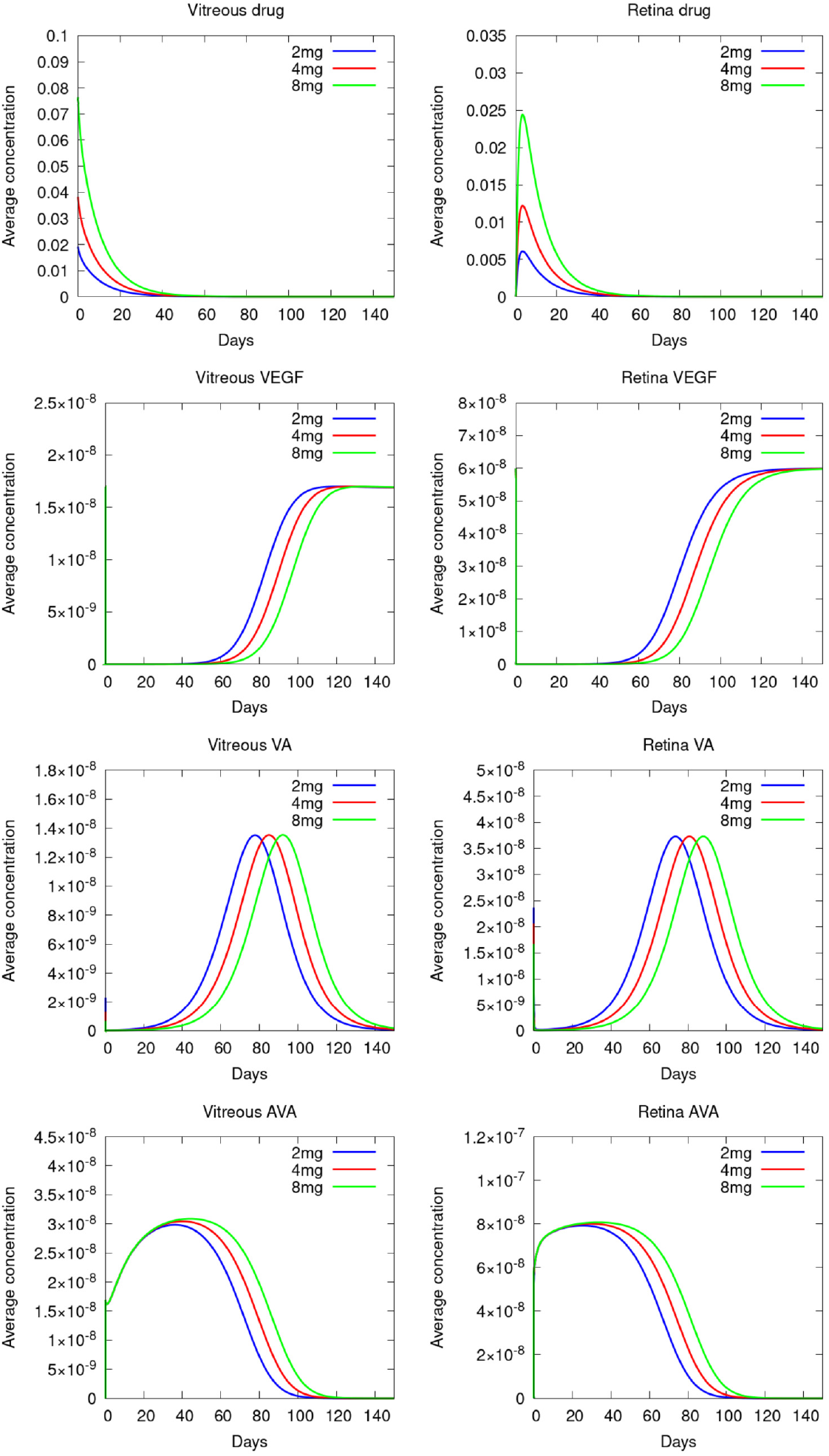
Comparison of the average concentrations over time in mol*/*m^3^ in the vitreous and the retina for different doses of an aflibercept injection.

In the future, the derived models and simulations could be used to further study different doses and intervalls between injections. Furthermore new developed drugs like brolucizumab and faricimab could be analysed and compared to aflibercept and ranibizumab.

## 3 Conclusion

In this work we derived a PDE model for the treatment of AMD. The model includes a fluid model for the vitreous using either a Burgers-type model for a healthy vitreous or the Navier-Stokes equations in case of a vitrectomy. The drug therapy model consists of a two-compartment model for the vitreous and retina coupled via interface conditions. In the vitreous we have four coupled convection-diffusion-reaction equations and in the retina four diffusion-reaction equations taking into account the binding processes of the drug and the VEGF. We derived the corresponding variational formulations and first discretized in time before using the finite element method for spatial discretization. Since the equations are nonlinear we derived the necessary Gateaux derivatives for Newton’s method.

First we showed numerical results for the fluid flow in the vitreous with a mean velocity of 7.25 · 10^−9^ m*/*s. The half-life time of the drugs were 5.6 for aflibercept and 4.5 days for ranibizumab. Furthermore the simulations showed the VEGF reducing effect of the drug for two common drugs, namely aflibercept and ranibizumab. The results showed that the efficacy of the drugs heavily depends on the measured binding parameters. Furthermore the results indicate the necessity for multiple injection which is in accordance to medical procedure where the injections occur every 30 days for a certain period of time. Furthermore we showed that different injection locations only have a minor impact on the long-term VEGF concentration. The analysis of the accumulated fluxes over time showed that for a uniform ranibizumab injection about 80 % of the drug clears through the hyaloid membrane and only about 20 % of the drug clears into the retina. Finally the simulations have given further insight into the binding and dissociation of the complexes over time as well as information about the VEGF concentration in the retina and the vitreous after an intravitreal injection.

## Data Availability

All relevant data are within the manuscript and its Supporting Information files.

## Acknowledgments

This work was embedded in the project BiTWerk of the University of Kassel and in the DFG Research Training Group 2749 Multiscale Clocks.

